# The Role of Self-efficacy Towards Health-promoting Lifestyle among Patients with Non-communicable Disease: A Systematic Literature Review

**DOI:** 10.1101/2025.01.05.25320027

**Authors:** Sharifah Maziah Syed Shamsuddin, Norfazilah Ahmad, Rosnah Sutan, Roszita Ibrahim

**Affiliations:** Department of Public Health Medicine, Faculty of Medicine, Universiti Kebangsaan Malaysia, Cheras, Kuala Lumpur, Malaysia

**Keywords:** Self-efficacy, Health-promoting lifestyle, non-communicable disease, NCD

## Abstract

**Background:** Non-communicable diseases (NCDs) will be a major cause of death worldwide by 2023. One effective strategy for preventing and managing NCDs is the implementation of health-promoting lifestyle intervention programs. Self-efficacy is a key factor associated with the adoption of health-promoting lifestyle practices. This review aims to examine the role of self-efficacy in fostering health-promoting lifestyles among patients with NCDs.

**Method:** A literature search was conducted across three scientific databases — Scopus, PubMed, and Web of Science —targeting original articles published in English between 2015 and 2024 that identified health-promoting lifestyle as an outcome. The quality of the eligible articles was assessed using the Joanna Briggs Institute (JBI) Critical Appraisal Tools, and the findings were synthesized through content analysis.

**Results:** The initial search identified 308 citations. A total of eight studies meeting the criteria of the JBI Critical Appraisal Tools were included, highlighting the direct effects of self-efficacy. Self-efficacy was demonstrated as a positive predictor of health-promoting lifestyle as a whole construct, as well as the physical activity dimension, among patients with NCDs, particularly those with hypertension, diabetes, cardiovascular disease, chronic obstructive pulmonary disease (COPD), and acute stages of cancer.

**Limitation:** All included studies were cross-sectional design. Therefore, the evidence quality was relatively low and exhibited a high risk of bias. Furthermore, there was language bias as only English-language publications were selected.

**Conclusion:** The findings of this review will guide healthcare providers in enhancing self-efficacy as a crucial positive predictor of health-promoting lifestyles among patients with non-communicable diseases. This approach can be integrated into clinic consultations and intervention programs. Future studies are warranted to evaluate the effectiveness of self-efficacy in improving a health-promoting lifestyle.

## Introduction

Non-communicable disease (NCD) is a chronic nature of diseases like cardiovascular diseases (stroke and myocardial infarction), cancers, diabetes, and chronic respiratory diseases (chronic obstructive pulmonary disease and asthma). NCD contributed to 74% of the cause of death worldwide (1). The WHO has established a Global Action Plan for the Prevention and Control of NCDs (2013–2020), which has been extended to 2030. This plan aims to reduce premature mortality from NCDs by one-third through effective prevention and treatment strategies, aligning with the Sustainable Development Goals (SDGs), particularly target 3.4, which focuses on reducing NCD-related deaths by 2030 (2).

The risk factors for NCD can be divided into non-modifiable and modifiable factors. The non-modifiable factors are age, gender, and family history. The modifiable factors related to practicing unhealthy lifestyles like physical inactivity, unhealthy diet, smoking, and harmful alcohol consumption (1, 3). Lifestyle is a way used by an individual and refers to the characteristics of a special time and place. It includes daily behaviour and functions of individuals in work, physical activities, happiness, and diet (4). One of the effective strategies in NCD prevention and management is through promoting a healthy lifestyle intervention program approach (5) which, has demonstrated a good impact on health outcomes (6), NCD prevention (7–9), reduces the risk of hypertension (10), and decreases financial burden on healthcare (11, 12).

The WHO promotes a health-promoting lifestyle by encouraging behaviour that reduces the risk factors associated with NCDs (2). Health-promoting lifestyle is defined as a multidimensional concept that is contributed by self-initiated action, various levels of behaviour, and self-perception by individuals to maintain or improve their wellness level, self-actualization, and individual fulfilment (13). This concept is based on behaviour, health belief (14), and Pender’s health-promotion models (13). Health-promoting lifestyle promotes health through lifestyle in six dimensions: i) health responsibility, ii) physical activity, iii) nutrition, iv) interpersonal relationships, v) spiritual growth, and vi) stress management (15, 16). Spiritual growth and physical activity demonstrated the highest and lowest scores of health-promoting lifestyle dimensions, respectively (17).

Studies have reported that many factors are associated with health-promoting lifestyle practices like socio-demographics (18, 19), illness perception (20), social support (21), and self-efficacy (9, 22–24). Self-efficacy is an individual’s belief in their ability to execute behaviour necessary to produce specific performance attainments (25). Self-efficacy influences patients’ confidence in their ability to adopt and maintain healthy lifestyle changes, which is crucial for managing NCDs. Self-efficacy is one of the Social Cognitive Theory (SCT) dimensions. The implementation of SCT towards NCD management and intervention in the community is increasingly recognized to enhance community health behaviour (26). Higher self-efficacy correlates with an increased likelihood of engaging and sustaining in health-promoting lifestyle activities, including physical activity and healthy nutrition (27). Hence, there is a need to understand the role of self-efficacy for healthcare providers to design targeted interventions that enhance patients’ confidence in their abilities to manage the NCDs.

There are various instruments available to measure self-efficacy through quantitative and qualitative approaches. Among quantitative instruments measuring self-efficacy include; the Self-Rated Abilities Scale for Health Practice (SRAHP) (9), Cancer Survivors’ Self-Efficacy Scale (CSSES) (28), General Self-Efficacy Scale (29), and Perceive Self-Efficacy Scale (PSES) (30). Qualitative instruments for measuring self-efficacy focus on exploring individuals’ perceptions and interpretations of their self-efficacy beliefs through open-ended questions, interviews, or think-aloud protocols (31, 32). Many studies conducted focusing on self-efficacy toward health-promoting lifestyles among different study populations. Therefore, this review aims to examine the role of self-efficacy in health-promoting lifestyles among patients with NCD and synthesize these findings in a single article.

## Methods

This review followed the Preferred Reporting Items for Systematic Reviews and Meta-Analysis (PRISMA) guidelines (33). The research question was developed using the PEO framework (P: population; E: exposure and O: outcome), focusing on the role of self-efficacy in a health-promoting lifestyle among patients with NCD. The population of this review is confined to the NCD population. The intervention for this study is the self-efficacy among the NCD patients and the outcome for this study is the health-promoting lifestyle among this study population.

### Search strategy

The comprehensive article search focused on the role of self-efficacy in health-promoting lifestyles among patients with NCD. The search for relevant articles was conducted across three indexed databases: Scopus, PubMed, and Web of Science, from 25^th^ to 28^th^ October 2024. Boolean operators combined keywords and synonyms related to the study population, intervention, and outcome. The terms were adapted to the specific requirements of each database to ensure comprehensive coverage. The search string was designed as shown in Table 1.

**Table 1.**
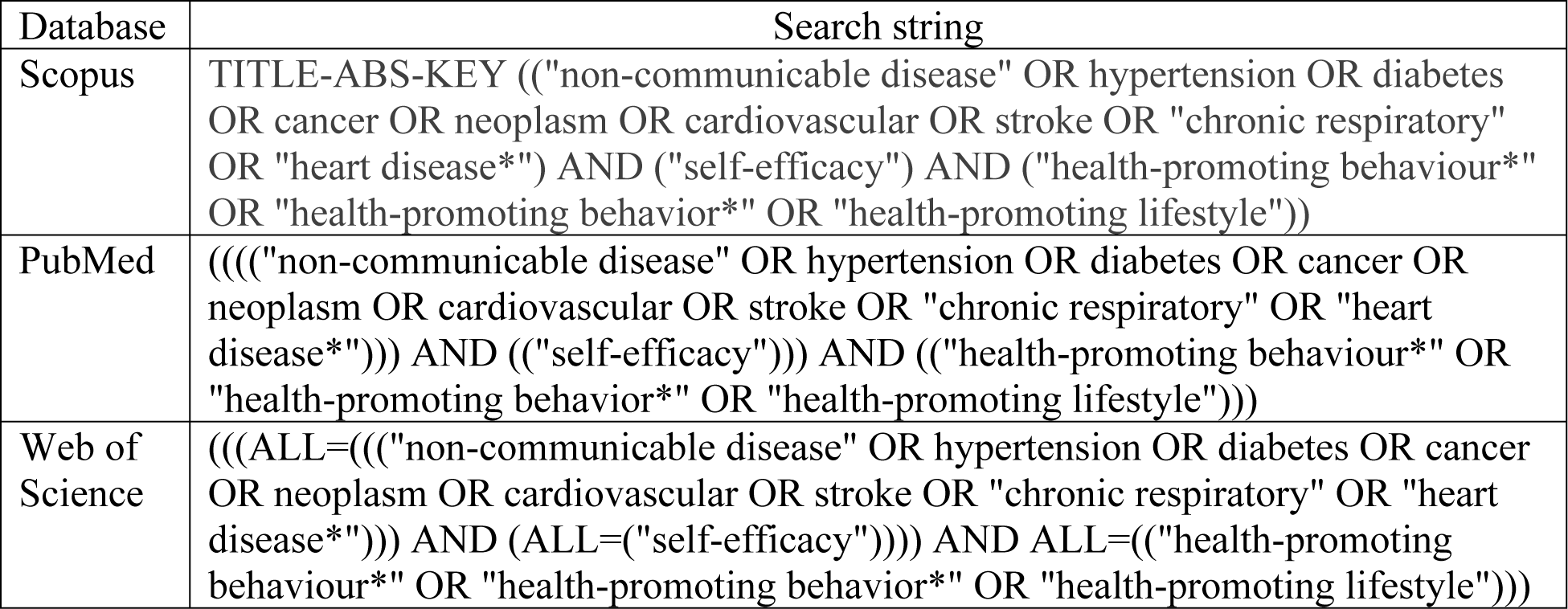
Keywords used in the screening process.

| Database | Search string |
| --- | --- |
| Scopus | TITLE-ABS-KEY (("non-communicable disease" OR hypertension OR diabetes OR cancer OR neoplasm OR cardiovascular OR stroke OR "chronic respiratory" OR "heart disease*") AND ("self-efficacy") AND ("health-promoting behaviour*" OR "health-promoting behavior*" OR "health-promoting lifestyle")) |
| PubMed | ((("non-communicable disease" OR hypertension OR diabetes OR cancer OR neoplasm OR cardiovascular OR stroke OR "chronic respiratory" OR "heart disease*")) AND (("self-efficacy"))) AND (("health-promoting behaviour*" OR "health-promoting behavior*" OR "health-promoting lifestyle")) |
| Web of Science | ((ALL=((("non-communicable disease" OR hypertension OR diabetes OR cancer OR neoplasm OR cardiovascular OR stroke OR "chronic respiratory" OR "heart disease*")) AND (ALL=("self-efficacy")))) AND ALL=(("health-promoting behaviour*" OR "health-promoting behavior*" OR "health-promoting lifestyle"))) |

### Inclusion and exclusion criteria

Two authors (SM and DNA) selected the eligible studies independently by reading the titles, abstracts, and full texts of the articles. The inclusion criteria were as follows: (i) Health-promoting lifestyle as the outcome; (ii) Publication in the English language; (iii) Original articles, and (iv) Publication from 2015 to 2024. We limited the publication date to this range (articles published in the past 10 years) to ensure that our review was based on recent literature. The exclusion criteria were as follows: (i) No factor of self-efficacy exposure; (ii) The terms “self-efficacy” and “health-promoting lifestyle” were not properly defined; and (iii) Articles focusing on populations other than NCD patients.

### Eligibility

The initial search identified 308 citations, of which 173 records were excluded based on year, language, and article type using an automated tool. After removing seven duplicate records, 128 unique records were subjected to title and abstract screening. During the screening, 120 articles were excluded. This left a total of eight articles for appraisal. Figure 1 presents the study selection process according to the PRISMA flow diagram.

**Figure 1.**
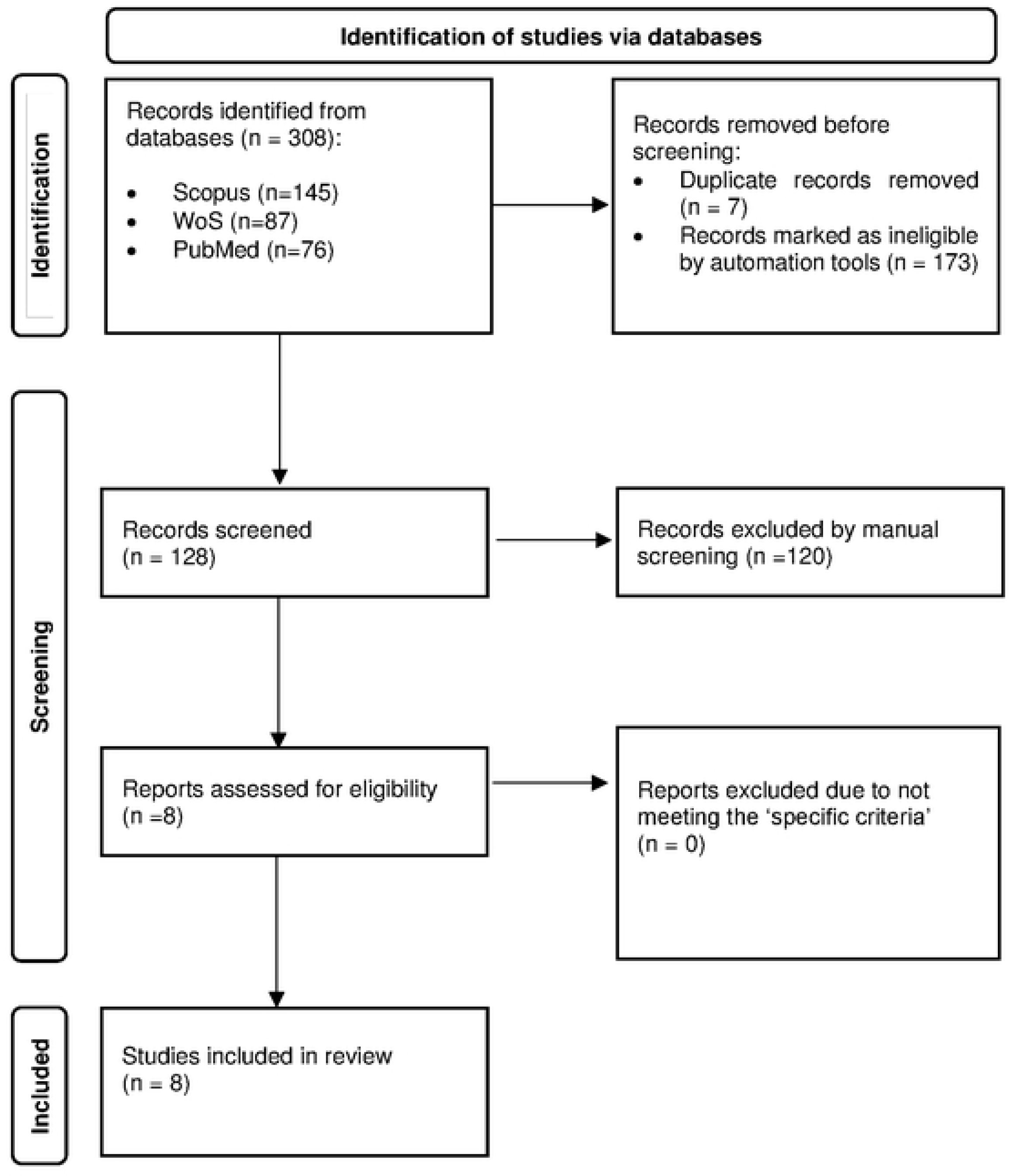
Study selection process according to the PRISMA flow diagram.

### Quality assessment of studies

Three authors (SM, RS, and DNA) critically appraised the quality of the included articles independently using the Joanna Briggs Institutes (JBI) Critical Appraisal Tools. The quality assessment was based on the study design checklist (34, 35) and the results are summarised in Table 2.

**Table 2.** Joanna Briggs Institutes Critical Appraisal Tools.

| Article No | Article | 1 | 2 | 3 | 4 | 5 | 6 | 7 | 8 | Overall (include /exclude) | Comment |
| --- | --- | --- | --- | --- | --- | --- | --- | --- | --- | --- | --- |
| 1. | Hsia et al. 2024 (30) | Y | Y | Y | Y | Y | Y | Y | Y | include | NA |
| 2. | Park et al. 2022 (28) | Y | Y | Y | Y | Y | Y | Y | Y | include | NA |
| 3. | Sebastian et al. 2021 (21) | Y | Y | Y | Y | Y | Y | Y | Y | include | NA |
| 4. | Posai et al. 2021 (36) | Y | Y | Y | Y | Y | Y | Y | Y | include | NA |
| 5. | Chantakeeree et al. 2021 (9) | Y | Y | Y | Y | Y | Y | Y | Y | include | NA |
| 6. | Hamed Bieyabanie & Mirghafourvand 2020 (29) | Y | Y | Y | Y | Y | Y | Y | Y | include | NA |
| 7. | Kim et al. 2020 (23) | Y | Y | Y | Y | Y | Y | Y | Y | include | NA |
| 8. | Giena et al. 2018 (22) | Y | Y | Y | Y | Y | Y | Y | Y | include | NA |

Y=yes, N=No, U=Unclear, NA= Not applicable

### Data extraction tool

Two authors (SM, RS, and DNA) extracted the data by screening abstracts and full texts separately. The extracted information included the author, publication year, study country, type of NCD, self-efficacy instrument, self-efficacy score, the role of self-efficacy, health-promoting lifestyle construct by total or dimension and instrument, and statistical analysis.

## Results

### Background of the eligible studies

Eight studies were included in this review (Table 2). Most of the studies were conducted in Thailand (n=2) and Korea (n=2), with the remaining studies conducted in China, India, Indonesia, and Iran. All articles were published between 2018 to 2024. A total of eight included studies were cross-sectional designs. The study’s population was patients with diabetes, hypertension, heart disease, cardiovascular disease, COPD, and cancer.

### The role of self-efficacy toward health-promoting lifestyle among patients with NCD

A total of eight studies demonstrated a significant positive relationship between self-efficacy and health-promoting lifestyle. Strong correlation was reported by two studies conducted in Thailand; i) among hospitalized patients with NCD during the second wave of COVID-19 (36) and ii) older persons with hypertension (9). Other studies reported that older persons with NCD in urban areas had higher self-efficacy and health-promoting lifestyles than those in rural areas (9, 37).

A moderate correlation was demonstrated by two studies conducted in Korea among thyroid cancer survivors (28) and patients with hypertension (23). Among thyroid cancer survivors, self-efficacy significantly affects the health-promoting lifestyle in the acute cancer stage (28). A recent study conducted in India among patients with diabetes also demonstrated a moderate correlation between self-efficacy and health-promoting lifestyle (21). A low positive correlation was reported by studies conducted in Iran and Indonesia among breast cancer survivors (29) and older patients with hypertension (22), respectively. Another study conducted among breast cancer survivors in China demonstrated a positive relationship between self-efficacy and health-promoting lifestyle (30).

Among all included studies, only three studies showed that self-efficacy was positively correlated with dimensions of health-promoting lifestyle. Two studies were conducted among breast cancer survivors (29, 30) and one study among patients with diabetes mellitus (21). The outcome analysis of this study focused only on the physical activity and nutrition dimension of a health-promoting lifestyle rather than the construct as a total (21).

### Self-efficacy instruments

A total of six types of questionnaires have been used to measure the self-efficacy variable: (i) SRAHP (9, 22, 30), (ii) Korean version Cancer Survivors’ Self-Efficacy Scale (CSSES-K) (28), (iii) General Self- Efficacy Scale (29), (iv) PSES (36), (v) Social Cognitive Theory (SCT) questionnaire (21), and (vi) Self-efficacy assessment tool for patients with hypertension (23). There are various self-efficacy scores demonstrated among different study populations using different self-efficacy measuring instruments as depicted in Table 2. A higher self-efficacy score indicates a higher level of self-efficacy (9).

**Table 2.**
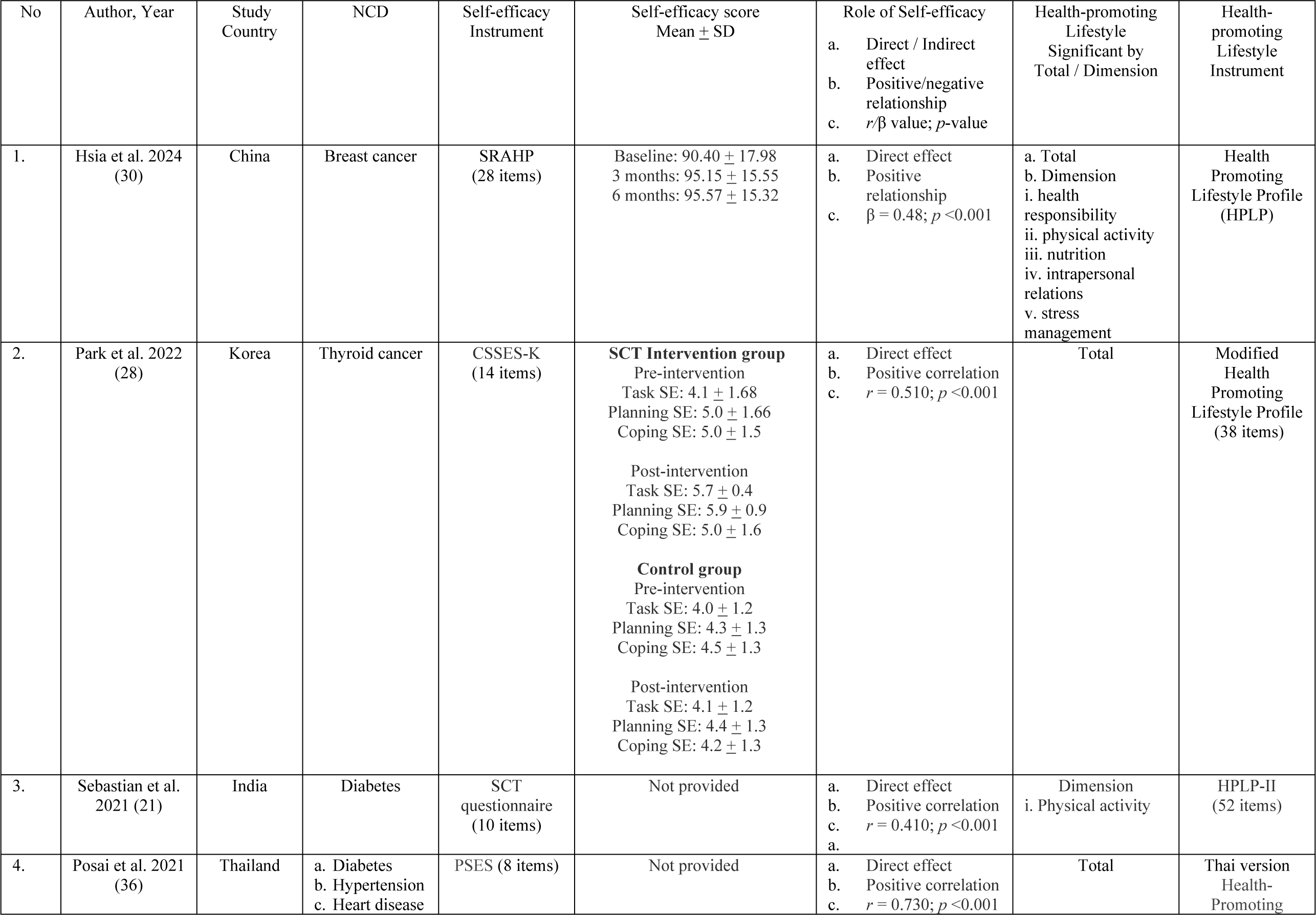
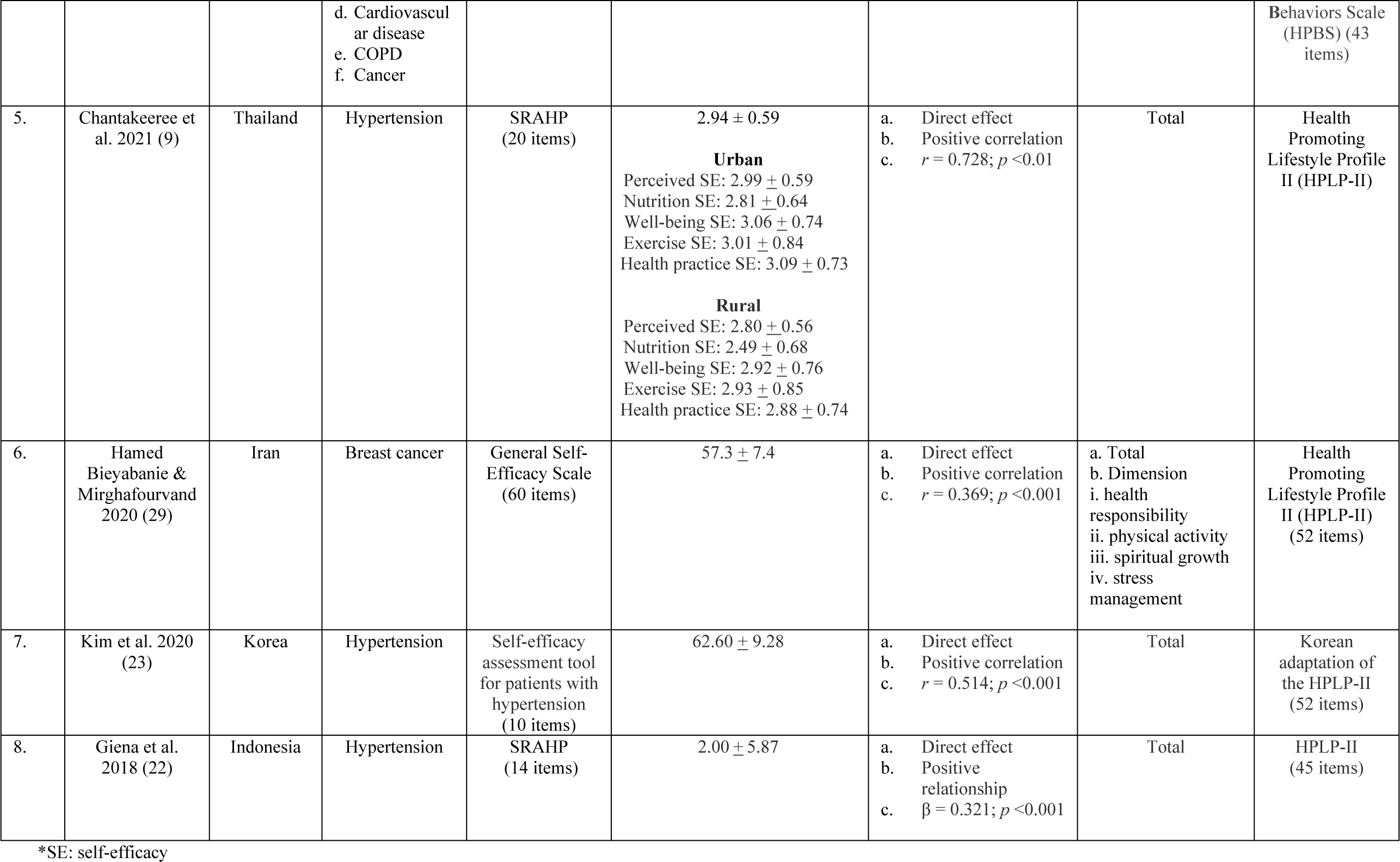
Characteristics of included studies on the role of self-efficacy.

## Discussion

Self-efficacy plays an important role in health-promoting lifestyles among cancer survivors by influencing the treatment aspects and recovery process. This includes coping strategies, physical activity, and psychological resilience (25). In view of coping with treatment aspects, high self-efficacy is associated with better coping strategies among cancer survivors (25). This allows cancer survivors to engage in a health-promoting lifestyle to manage distress regarding treatment side effects and adhere to treatment regimes effectively (25, 38).

Physical activity is an important health-promoting lifestyle dimension for cancer survivors and diabetic and hypertensive patients. It can alleviate treatment side effects and enhance quality of life among cancer survivors, diabetic, and hypertensive patients. Regular physical activity like brisk walking, cardiac rehabilitation, and cycling is vital in patients with NCD. Self-efficacy is a strong predictor of physical activity adherence among patients with NCD (39). Self-efficacy helps patients with NCD maintain their physical activity levels despite having barriers like fatigue and cancer treatment challenges (25).

Self-efficacy influences the health-promoting lifestyle in short- and long-term maintenance. Studies reported patients with NCD who possess a high self-efficacy correlate with better adherence to prescribed medication regimes. This is important in hypertension and diabetic control. Patients with high self-efficacy are more proactive in the health responsibility dimension such as regular self-performing blood glucose (40) and pressure monitoring (41). The nutrition dimension of a health-promoting lifestyle is comprised of a low-sodium and low-glucose diet which is critical for blood pressure and glucose control, respectively. Patients with high self-efficacy feel more confident about making dietary changes and compliance (41).

Higher self-efficacy indicates a higher level of interpersonal relations and spiritual growth. Expansion of interpersonal relations will enhance patients’ confidence in discussing their symptoms and disease treatment plans. This will give rise to a productive discussion on the personalised management of pharmacotherapy, rehabilitation, and self-care (42). Developing personalised treatment plan required high spiritual growth to initiate, practice, and sustain the behaviour changes. High spiritual growth leads patients to perceive that they are capable of managing their disease resulting in attending empowerment programs like cardiac rehabilitation and consultation clinics that finally result in good disease control (14).

Self-efficacy levels can be increased through a patient empowerment program, motivational interviewing techniques, peer support groups, and engaging family members in the NCD care process. Studies demonstrated that by further emphasizing supportive family environments, self-efficacy among patients with NCDs can be increased (43, 44). Family support assists in emotional, and practical, and boosts patient’s confidence in managing their NCDs (40, 45). These approaches can be implemented in primary and secondary healthcare facilities by integrating family support in NCD management. Personalised management can be tailored to the specific patient for health-promoting lifestyle improvement.

Another personalised management approach to enhance self-efficacy among patients with NCD is through the motivational interviewing technique. Motivational interviewing is a patient-centered counseling technique to enhance intrinsic motivation for change by exploring and resolving barriers. This approach respects the autonomy of the patient, allowing them to take ownership of their behavioural change process. The technique seeks to draw out the patient’s motivations for change rather than imposing external motivations. This is crucial for building self-efficacy, as patients are more likely to commit to changes that they feel personally motivated to pursue.

## Strengths and limitations

All of the included studies were cross-sectional design, which directed this systematic review to a high risk of bias and the inability to report causal relationships. Restriction on English-language articles limits the range of article inclusion, which may exclude good-quality articles published in other languages. The use of tools for translation can lead to an increased risk of instrument bias. Nevertheless, the search strategy has resulted in literature sources from various countries where English is not the first language (China, Korea, Iran, and Indonesia).

The strength of this study is the eligibility criteria that only include a clear definition of self-efficacy toward a health-promoting lifestyle. Additionally, to the best of our knowledge, this is the first systematic review of synthetizing research evidence on the role of self-efficacy in health-promoting lifestyles among NCD patients. This systematic review highlights the significance of the self-efficacy approach in improving health-promoting lifestyles among various population backgrounds.

## Conclusion

In conclusion, self-efficacy is a vital positive predictor of a health-promoting lifestyle as a construct or by its dimension. Self-efficacy levels can be increased through individual and community program approaches. These approaches can be implemented in primary, secondary, and tertiary healthcare facilities by tailoring to the specific population background to reduce the global NCDs’ burden.

## Data Availability

All relevant data are within the manuscript and its Supporting Information files.

## Acknowledgments

The authors would like to thank the Dean of Medical Faculty (FF 2024–129) and Department of Public Health Medicine, Faculty of Medicine, Universiti Kebangsaan Malaysia for supporting this review and the Director General of Health Malaysia for his permission to publish this article.

## Supporting information

S1 Table. Table 1. The PRISMA 2020 checklist.

S2 Table. Table 2. The PRISMA 2020 Abstract checklist.

S3 Table. Table 3. Studies identified in literature search.

S4 Table. Table 4. The detailed description of studies included in systematic literature review.

Fig.1. Figure 1. Study selection process according to PRISMA flowchart.

S1 Appendix. Conflict of Interests Statement and Author Contributions

## Notes

### Competing Interest Statement

The authors have declared no competing interest.

### Funding Statement

The author(s) received no specific funding for this work.

